# Precision medicine for developmental and epileptic encephalopathies in Africa – strategies for a resource-limited setting

**DOI:** 10.1101/2022.08.17.22278768

**Authors:** Alina I. Esterhuizen, Nicki Tiffin, Gillian Riordan, Marie Wessels, Richard J. Burman, Miriam C. Aziz, Jeffrey D. Calhoun, Jonathan Gunti, Ezra E. Amiri, Aishwarya Ramamurthy, Michael J. Bamshad, Deborah A. Nickerson, Heather C. Mefford, Raj Ramesar, Jo M. Wilmshurst, Gemma L. Carvill

**Affiliations:** The South African MRC/UCT Genomic and Precision Medicine Research Unit, Division of Human Genetics, Institute of Infectious Diseases and Molecular Medicine, Department of Pathology, University of Cape Town, Cape Town, South Africa; National Health Laboratory Service, Groote Schuur Hospital, Cape Town, South Africa; South African National Bioinformatics Institute, University of the Western Cape, South Africa; Department of Paediatric Neurology, Red Cross War Memorial Children’s Hospital, Neuroscience Institute, University of Cape Town, South Africa; Division of Clinical Neurology, Nuffield Department of Clinical Neurosciences, John Radcliffe Hospital, University of Oxford, United Kingdom; Ken and Ruth Davee Department of Neurology, Northwestern University Feinberg School of Medicine, Chicago, IL; University of Washington Centre for Mendelian Genomics (UW-CMG); Department of Pediatrics, University of Washington, Seattle, WA; Department of Genome Sciences, University of Washington, Seattle, WA; Brotman-Baty Institute, Seattle WA; Centre for Paediatric Neurological Disease Research, St. Jude Children’s Research Hospital; Department of Pharmacology, Northwestern University Feinberg School of Medicine, Chicago, IL; Department of Pediatrics, Northwestern University Feinberg School of Medicine, Chicago, IL

**Keywords:** Epilepsy, Seizures, DEE, LMICs, genetic epilepsy, South Africa, genetic testing

## Abstract

**Purpose:** Sub-Saharan Africa bears the highest burden of epilepsy worldwide. A presumed proportion is genetic, but this aetiology is buried under the burden of infections and perinatal insults, in a setting of limited awareness and few options for testing. Children with developmental and epileptic encephalopathies (DEEs), are most severely affected by this diagnostic gap in Africa, as the rate of actionable findings is highest in DEE-associated genes.

**Methods:** We tested 235 genetically naïve South African children diagnosed with/possible DEE, using gene panels, exome sequencing and chromosomal microarray. Statistical comparison of electroclinical features in children with and without candidate variants was performed to identify characteristics most likely predictive of a positive genetic finding.

**Results:** Of 41/235 children with likely/pathogenic variants, 26/235 had variants supporting precision therapy. Multivariate regression modelling highlighted neonatal or infantile-onset seizures and movement abnormalities as predictive of a positive genetic finding. We used this, coupled with an emphasis on precision medicine outcomes, to propose the pragmatic “Think-Genetics” strategy for early recognition of a possible genetic aetiology.

**Conclusion:** Our findings emphasise the importance of an early genetic diagnosis in DEE. We designed the “Think-Genetics” strategy for early recognition, appropriate interim management and genetic testing for DEE in resource-constrained settings.

## INTRODUCTION

Epilepsy is one of the most common neurological conditions affecting approximately 50 million people worldwide(1). Whilst most epilepsy research is conducted in resource-equipped countries, the highest burden of the disease is carried by Sub-Saharan Africa (SSA), ascribed to the high rate of infections, perinatal insults, traumatic brain injury (TBI), as well as the under-resourced healthcare systems(2). The stigma and misconceptions surrounding epilepsy in some communities often prevent the individuals and caregivers from seeking medical help. The resulting economical and psychosocial burden on the people and families living with epilepsy in low and middle income countries (LMICs) demands improved understanding and interventions. This public health imperative has been recognised through the development of the Intersectoral Global Action Plan on Epilepsy and Other Neurological Disorders. Among its global targets is a 50% increase in epilepsy service coverage by 2031 (from that in 2021), and development of legislation promoting and protecting the human rights of people with epilepsy, in 80% of the member countries(3).

A sizable proportion of epilepsy in Africa is presumed to be genetic, but the genetic architecture is largely undetermined within the context of minimal research and limited clinical testing. Genetic epilepsies and the associated syndromes are frequently missed or misdiagnosed, and inappropriately treated(4–6). The consequences are especially dire for children with developmental and epileptic encephalopathies (DEEs), where early diagnosis and appropriate treatment are critical in mitigating the detrimental effects of ongoing seizures on the developing brain(7). On the opposite end of the economic spectrum in high income countries (HICs), next generation sequencing (NGS) gene panels, exome sequencing (ES) and chromosomal microarrays (CMA) are a routine part of diagnostic laboratory protocols, with genome sequencing (GS) having made the transition from research into the clinical laboratories in HICs(8). Testing is mainly focused on the DEEs, where the rate of actionable findings is highest(9, 10). New knowledge of epilepsy-associated genes, genotype-phenotype correlations and precision treatment approaches is swiftly translated into clinical practice(11–14).

These economic disparities highlight not only the gap in service provision but also the lacking genetic epilepsy research in Africa(5). Here, we describe the genetic architecture of early-onset epilepsies in South African (SA) patients, using gene panels, ES and CMA. Moreover, our detailed analysis of the electroclinical characteristics in individuals with and without a detected genetic cause, identified features which may be predictive of a positive genetic finding. We use this, coupled with an emphasis on actionable genes, to propose a pragmatic strategy for early recognition, testing and precision treatment for DEEs in LMICs. We suggest that this approach, whilst different from that followed in HICs, may be effective in bridging the disparities in diagnosing genetic epilepsies in LMICs.

## MATERIALS AND METHODS

### Recruitment site

Most study participants were recruited from the Epilepsy Clinic at the Red Cross War Memorial Children’s Hospital (RCWMCH) in Cape Town, a tertiary teaching hospital affiliated to the University of Cape Town (UCT) and the main specialist care centre for paediatric epilepsy in SSA. Patients present either directly to the hospital, or via specialist referrals for drug-resistant epilepsy assessments. The neurology service has a dedicated paediatric neurophysiology unit with access to video EEG telemetry and invasive monitoring by trained neurophysiology staff and accredited paediatric neurologists. All children with recurrent seizure onset under two years of age undergo assessments for aetiology indicators and seizure semiology and managed for ongoing care. Standard assessments include exclusion of metabolic (urinary organic and amino acids, plasma ammonia, biochemistry, liver function and cerebrospinal fluid protein levels as well as glucose and lactate paired with plasma) and structural pathology (CT scan acutely, then MRI brain as part of initial epilepsy assessment and typically by 2 years of age for optimal myelination), as well as assessment for immune-mediated encephalitis, where indicated. The service has access to ancillary services inclusive of rehabilitation and child development. Whilst there is capability of screening for most seizure aetiologies, access to genetic analysis is currently lacking.

### Study population

We recruited 235 genetically untested SA children between 2015 and 2019, with early-onset, drug-resistant epilepsy, and a diagnosis or suspicion of DEE, with no known infectious, metabolic, immune, structural (non-genetic) or another acquired cause. The age of onset was extended to eight years, to ensure inclusion of possible late-presenting DEEs. A small number of children were recruited by affiliated neurologists in private practice. The study group comprised of 121 males and 113 females, self-reported as Indigenous Black African (*n* = 102); Mixed Ancestry (*n* = 91), Asian (*n* = 1), European (*n* = 17) and Other (*n* = 23), broadly representing the population demographic of the SA Western Cape region. The group included 22 children clinically diagnosed with Dravet syndrome (DS), who were investigated in a pilot project(15).

### Clinical Information Collection and Assessment

Information was collected by clinical assessment, review of patient records, parent/guardian interview and/or clinician questionnaire, and captured in a custom-designed RedCap database(16) (Suppl. Table 1, Suppl. Note 1). To maintain consistency, the data was captured by a team of the directly involved clinicians and the molecular geneticist. Data which could not be confidently documented was preferentially excluded.

### Genetic testing

DNA was extracted from peripheral blood using standard methods.

#### DEE-associated gene panel

All patients were tested with a panel of 71 DEE-associated genes, using previously described methods(17–19) (Suppl. Note 2). Variants were filtered and prioritised for *de novo* and recessive variants. Only nonsynonymous, splice-site and frameshift changes were assessed. Segregation analysis was performed where parental samples were available. Exon/gene level copy number calling was performed as previously described, and findings confirmed using multiple ligase-dependent probe amplification (MLPA) or CMA(20). Variant classification followed published guidelines(21). Microsatellite analysis was performed to confirm biological relationships.

#### CMA for genome-wide copy number variant (CNV) detection

Seventy eight patients with no findings on the gene panel were selected for CMA analysis with a custom 4 × 180K Comparative Genomic Hybridisation array (Agilent) (Suppl. Note 3). Patient selection was based primarily on the availability of high-quality DNA. ClinGen CNV Pathogenicity Calculator was used for CNV interpretation according to the published scoring metrics(22).

#### Trio ES

Twenty patient/parent trios were selected for ES, based on the absence of informative findings on the gene panel or CMA, and availability of parental DNA. ES was performed as previously described, using the Illumina HiSeq 2000using platform and the VCRome v.2.1 target-capture reagents (Roche Nimblegen)(23). We prioritised *de novo* and recessive variants for further pathogenicity assessment. Positive missense-z and pLi scores were used as a criterion for non-truncating variant filtering in candidate genes(24).

### Clinical correlation

The available electroclinical information and response to ASMs in each patient with P/LP variant was carefully compared with that described for the gene/variant, to finalise the diagnosis and determine appropriateness of treatment (e.g., avoid sodium channels blockers in *SCN1A*-DS). The functional effect (if known), of the variant was used to further refine options for precision treatment (especially relevant to the ion channel genes)(11).

### Statistical analysis

The clinical characteristics assessed in the study population were described with descriptive statistics using R software (v. 4.1.1) and the R Studio interface (25). Comparisons were made between the characteristics in: (1) patients with candidate variants (any class) vs those without; (2) patients with candidate SNV/indels vs those without; (3) patients with candidate CNVs and those without. In addition, multiple linear regression modelling was used to assess the association between selected clinical features (variables) and having an identified candidate variant. The variable selection was based on (1) statistical significance obtained in the preliminary data comparison (p-value <0.5), (2) completeness of the data set (Redcap entry for most patients), and (3) obtainability through clinical examination.

## RESULTS

### Patient Demographic and Phenotypes

Of the 235 probands, 77 were recruited as singletons (probands), 108 proband/mother duos and 50 parent/proband trios. At least one episode of status epilepticus was recorded in 84/235 (36%) children. Global developmental delay (GDD) prior to seizure onset was recorded in 58/235(25%) cases. Abnormal movements were noted in 39/235(17%), and dysmorphic features in 26/235(11%) cases. Autism spectrum disorder (ASD) was diagnosed in 30/235(13%) children and attention difficulties in 21/235(9%) (Suppl. Tables 3, 4 and 5). The overall median age at seizure onset was 8 months (IQR: 3,18). Neonatal onset (0 - 1 month) was noted in 19 cases, infantile onset (2 – 24 months) in 165, childhood onset (>24 months) in 33, and was unknown in 17 children. The median time between the seizure onset and seeking medical assistance was relatively short for the neonates and infants (one week and three months, respectively) but increased to nine months in the childhood-onset group (Table 1).

**Table 1.**
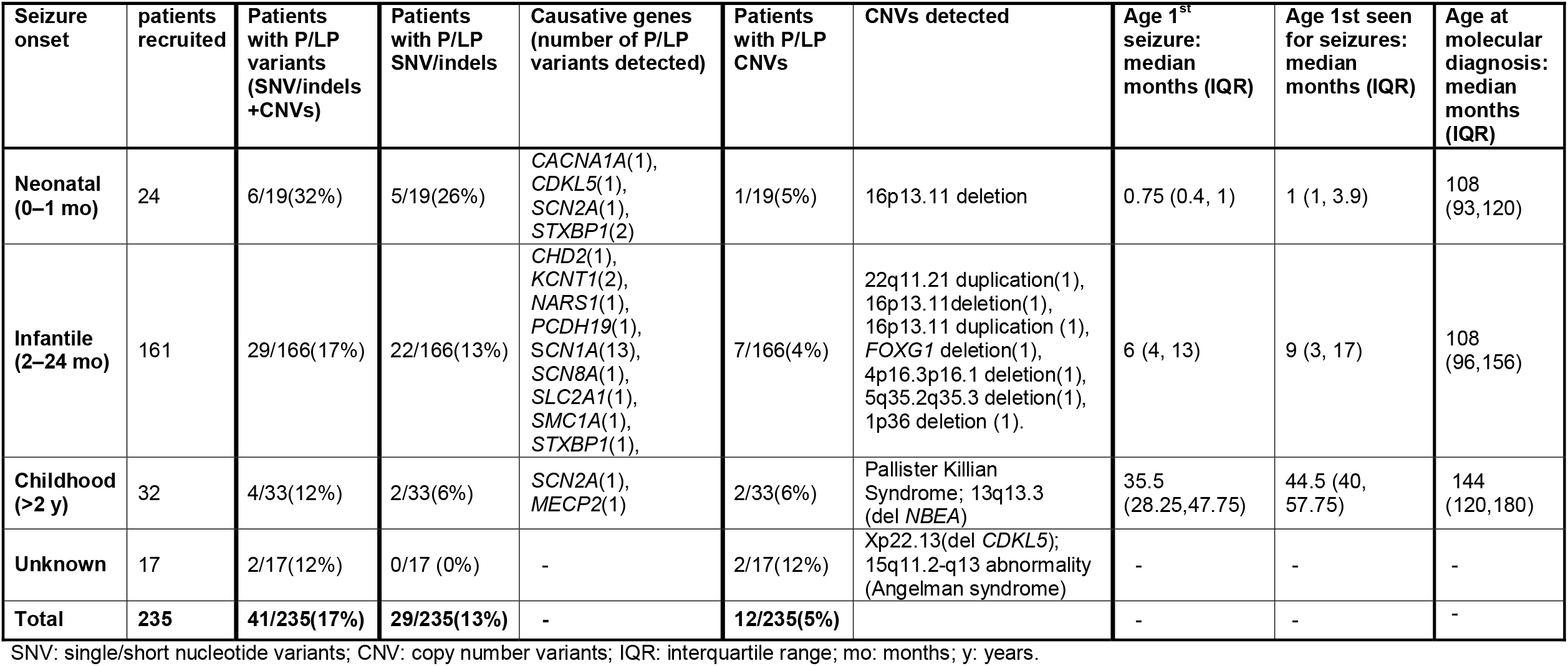
Ages at seizure onset and molecular diagnosis, and the causative genes in the children with P/LP variants.

During the course of the study, 14/231(0.06%), children were diagnosed with DEE due to other causes: acquired structural or infectious (*n* = 7), primary generalised dystonia (*n* = 1), tuberous sclerosis complex (TSC, *n* = 2), biotinidase deficiency (*n* = 1), thiamine deficiency (*n* = 1), Sturge Weber Syndrome (*n* = 1) and Moya-Moya disease (*n* = 1). We decided not to exclude these patients from the subsequent statistical analyses, as they form part of a realistic referral base of the state specialist epilepsy service in SA.

### The genetic architecture of early-onset epilepsy in South African children

Overall, rare genetic variants were detected in 52 patients: (P/LP) variants were identified in 41/235 (17%) children and 16 variants of uncertain significance (VUS) were detected in 12 (Suppl. Table 2)

#### Sequence Variants Detected with a Panel of 71 DEE-associated Genes

P/LP single/short nucleotide variants (SNV/indels) were identified in 28/235 patients (12%), in 12 DEE-associated genes. Segregation analysis revealed *de novo* occurrence in 10 cases. Complete patient/parent trios were not available for the remaining 19 cases, but the available parents tested negative for the putative variants. *SCN1A* was the highest yielding gene (*n* = 13), followed by *STXBP1* (*n* = 3), *SCN2A* (*n* = 2), *KCNT1* (*n* = 2), and one variant respectively in *CACNA1A, CDKL5, CHD2, MECP2, PCDH19, SLC2A1, SCN8A* and *SMC1A*. Five VUS were detected in *ATP1A2, KCNA2, SCN1A, SCN2A, and SCN3A*, respectively (Suppl. Table 2). More than two thirds of the detected P/LP variants were found in the ion channel genes(20). All P/LP *SCN1A* variants were in patients with a clinical diagnosis of Dravet Syndrome (DS) (*n* = 13) (Suppl. Table 1).

#### Genomic CNVs Detected with CMA

P/LP CNVs were identified in 12/78 (15%) patients, with microdeletion/duplication syndromes were identified and clinically confirmed in six: 1p36 deletion syndrome (*n* = 1), Wolf Hirschhorn syndrome (*n* = 1) Sotos syndrome (*n* = 1), 22q11.2 microduplication syndrome (*n* = 1), Angelman syndrome (*n* = 1) and Pallister-Killian mosaic syndrome (*n* = 1). Pathogenic whole-gene deletions were observed in three patients: a heterozygous *FOXG1* deletion in a child whose phenotype matched that of *FOXG1* deletion syndrome (26), a *CDKL5* gene deletion in a female whose phenotype was consistent with *CDKL5* Deficiency Disorder(27), and a deletion of *NBEA* at 13q13.3 in a child subsequently lost to follow up, hence, no clinical correlation was possible(28). Heterozygous loss at the 16p13.11 susceptibility region was detected in two patients, of which one was maternally inherited. Another patient had a maternally inherited 16p13.11 gain, as well as a novel *KCNA2* missense VUS. The *KCN2A* variant was not found in his mother (who tested positive for the 16p13.11 gain) but no paternal sample was available to establish/exclude *de novo* occurrence (Suppl. Tables 1 and 2).

#### ES in Parent/Child Trios reveals rare genetic causes and novel candidate genes for the DEEs

*De novo* analysis identified candidates in five genes: *ANGPT1* (*n* = 1), *COBLL1* (*n* = 1), *GLUL* (*n* = 1), *NARS* (*n* = 1) and *PLPPR4* (*n* = 1) (Suppl. Table 2). Of these, only the *de novo* recurrent *NARS* variant fulfilled criteria for pathogenicity and the patient’s clinical features aligned with the *NARS* neurodevelopmental phenotype described by Manole et al., (2020)(29). Analysis for autosomal recessive (AR) inheritance revealed compound heterozygosity for VUS in three genes (*UNC80, CELSR2* and *APC2*) (Suppl. Table 2). To our knowledge, only the *NARS* and *UNC80* genes have been definitively linked with phenotypes involving epilepsy. We engaged the GeneMatcher platform to connect with other investigators and learn more about the VUS detected in the other genes (30). None have been resolved to date.

### A genetic diagnosis impacts clinical care and management

Actionable variants were detected in eight genes (*CDKL5, KCNT1, PCDH19, SCN1A, SCN2A, SCN8A, SLC2A1 and STXBP1*)(9), supporting precision therapy for 63% (26/41) of the individuals with P/LP variants, and 11% (26/235) of the children overall(9) (Suppl. Table 2). ASM changes included dose optimisation (e.g., carbamazepine) or revisiting motivation for access to restricted ASMs (e.g., stiripentol). Knowledge of the genetic aetiology also enabled a more targeted multidisciplinary team (MDT) care and better insights into prognosis, disease course and complications (Suppl. Table 1, Figure 1). Cascade testing of the at-risk or affected relatives was offered to the families of the patients with maternally inherited CNVs (22q11.21 gain, 16p13.11 loss and 16p13.11 gain). In two of these cases, the mothers were mildly affected, which was only noted after testing the children. Prenatal analysis is now available to these mothers for potential future pregnancies.

**Figure 1.**
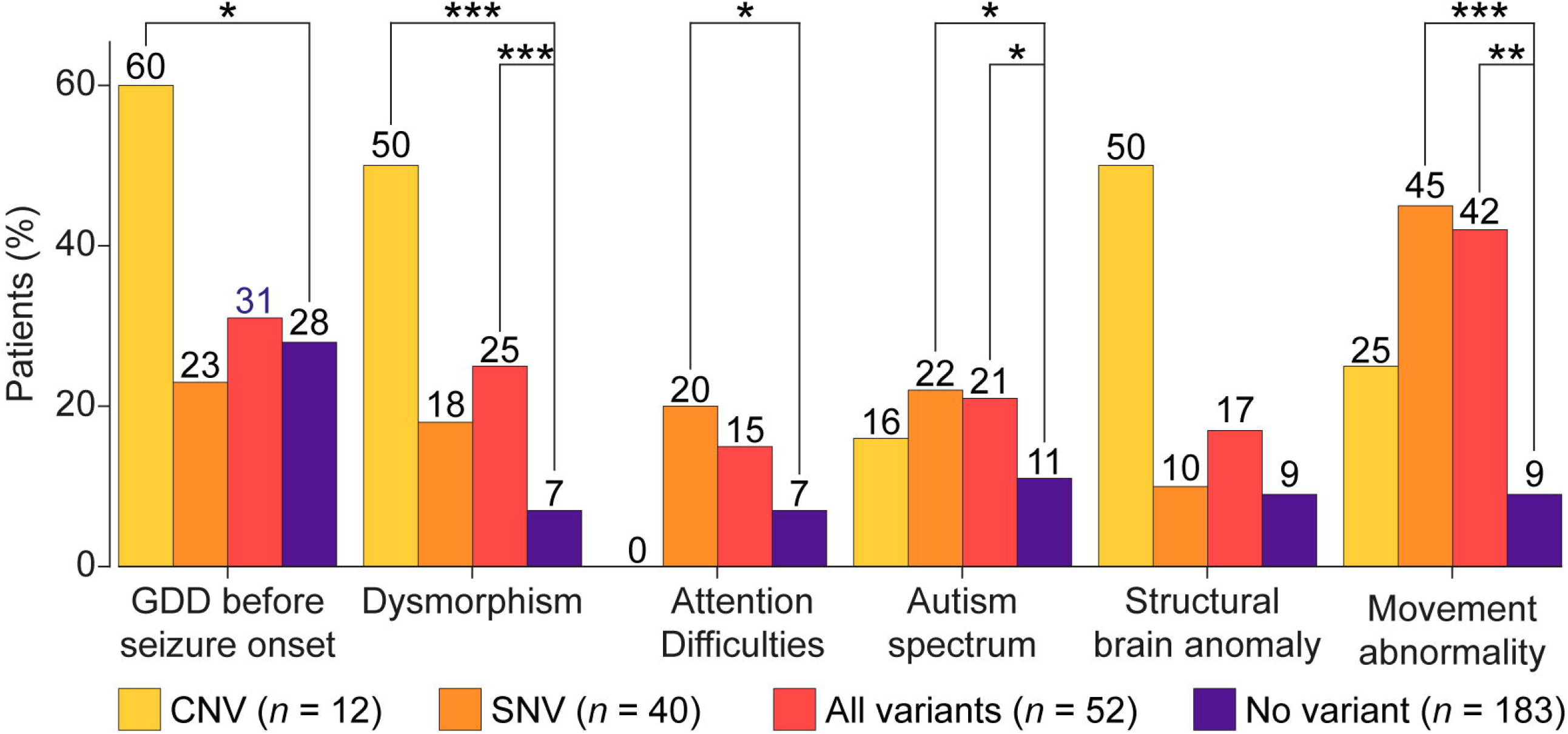
Case Box descriptions of three patients with *SCN2A* variants with different variant types, ages of seizure onset and responses to treatment, demonstrating the relevance of the functional effect of the underlying variant to precision treatment.

### Statistical analysis reveals specific clinical features more frequently associated with disease-causing SNV/indels and CNVs

Causative variants were identified, and clinical syndromes confirmed in 41/235(17%) children in our study. Of these, the mean age at a molecular diagnosis was 108 months (IQR: 93,120) in children with neonatal-onset epilepsy, 108 months (IQR:96,156) in the infantile-onset group and 144 months (IQR: 120,180) in the childhood-onset group (Table 1). Patients with neonatal-onset epilepsy had the highest proportion of P/LP SNVs/indels (32%, 6/19), followed by the infantile-onset (17%, 29/166) and the childhood-onset group (12%, 4/33). CNVs which were detected in approximately 5% of the children in each age-of-onset group (Table 1).

There were no significant differences between the seizure types recorded in individuals with and without P/LP variants, stratified per age at seizure onset (neonatal, infantile and childhood). The three most frequently noted seizure types in all groups were generalised tonic-clonic (GTC), focal and myoclonic seizures. Febrile seizures were prevalent among children with infantile-onset epilepsy, almost all diagnosed with DS (Suppl. Figure 1). Dysmorphism and GDD prior to seizure onset were noted in more than half (60%) of the patients with CNVs, compared to less than a third of patients with SNVs or no candidate variants. ASD and attention difficulties featured prominently among individuals with SNV/indels (20%). The SNV/indel group also had the greatest proportion of patients with movement abnormalities (45%) and the youngest age at first seizure (median 5 months (IQR: 2.9)). Structural brain anomalies were prominent in the CNV group (Figure 2, Suppl. Tables 3, 4 and 5).

**Figure 2.**
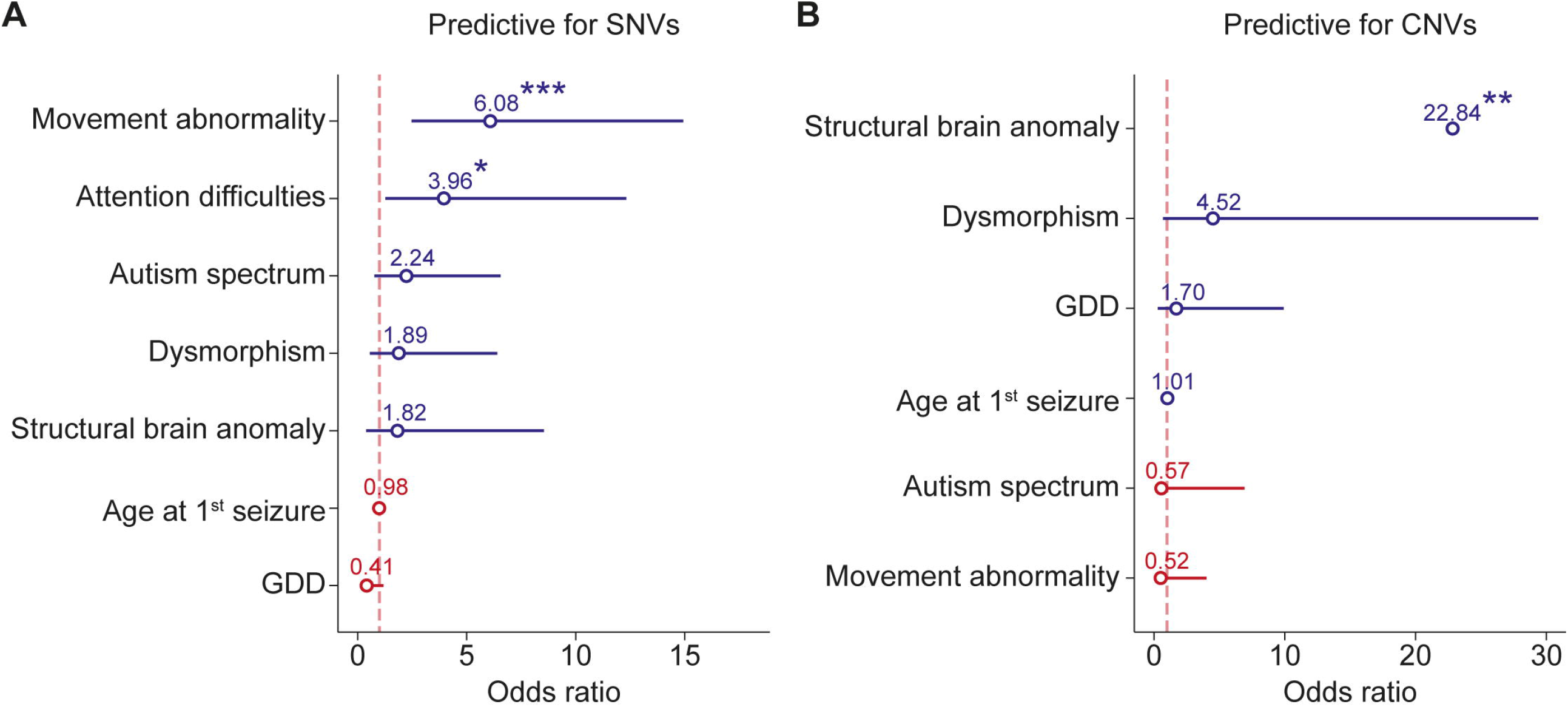
Statistical summary of selected clinical features in patients with candidate genetic variants. The number of patients with a variant in each class (i.e., CNV, SNV/indel, CNV/SNV/indel combined,) and the specific clinical feature are shown. For instance, 60% of the patients with a detected CNV had GDD before seizure onset, 23% of the patients with a detected SNV/indel had GDD before seizure onset, etc. No line/star: *p* >0.05; \**p* ≤ 0.05; \*\**p* ≤ 0.01; \*\*\**p* ≤ 0.001.

Multivariate logistic regression analysis suggested associations between movement abnormalities (OR: 6.68, 95%CI: 2.49, 15.2) and attention difficulties (OR: 3.96, 95%CI: 1.24, 12.3), with the presence of a candidate SNV/indel (Figure 3, model A; Suppl. Table 6), whereas structural brain anomalies achieved significance among patients with CNVs (OR: 22.84, 95%CI: 3.25, 181) (Figure 3, model B; Suppl. Table 7). These features were also noted in the combined variant model (SNV/indels and CNVs) (Suppl. Figure 2, Suppl. Table 8). ASD, dysmorphic features, GDD and age at first seizure did not show significant associations with the presence of a candidate variant in the multivariate regression model but did achieve statistical significance in the initial statistical analysis (*p*-value <0.05) (Figure 2, Suppl. Tables 3, 4 and 5).

**Figure 3.**
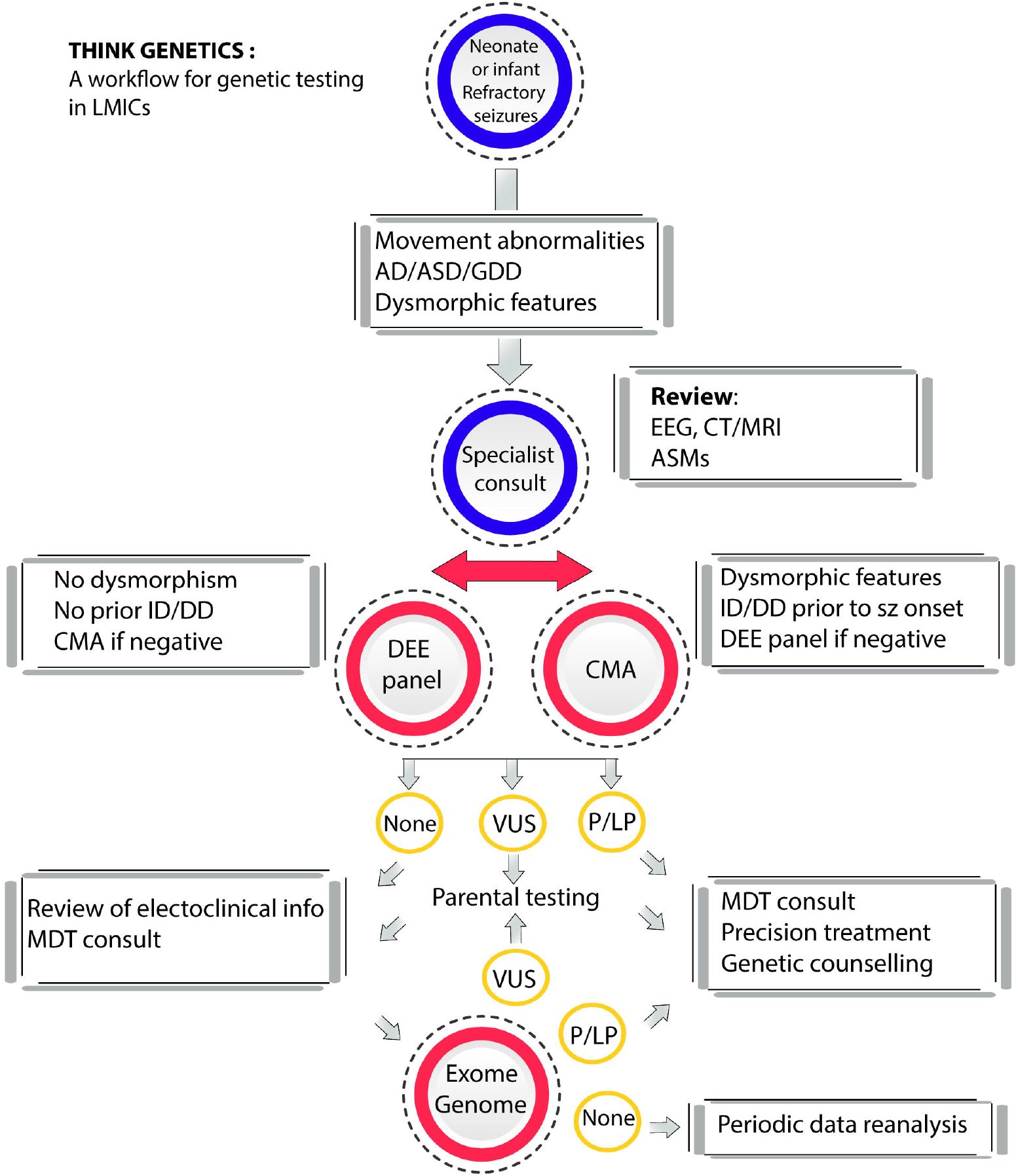
Multivariate Logistic Regression Analysis for assessment of association between selected clinical features in patients with and without an identified genetic aetiology. (A) SNV/indels: significant associations were observed with movement abnormalities and attention difficulties (B) CNVs: significant associations were observed with structural brain anomalies. \**p*-value <0.05 and \*\**p*-value <0.01

### A “Think-Genetics” decision tree for an early diagnosis of a suspected genetic epilepsy/DEE in the LMICs

We used three facets of our results to develop the Think-Genetics (TG) decision tree, designed to help clinicians at patient-entry in resource-constrained settings to recognise a possible genetic epilepsy/DEE, and initiate a process of MDT consultation and genetic testing (Figure 4):

**Figure 4.**
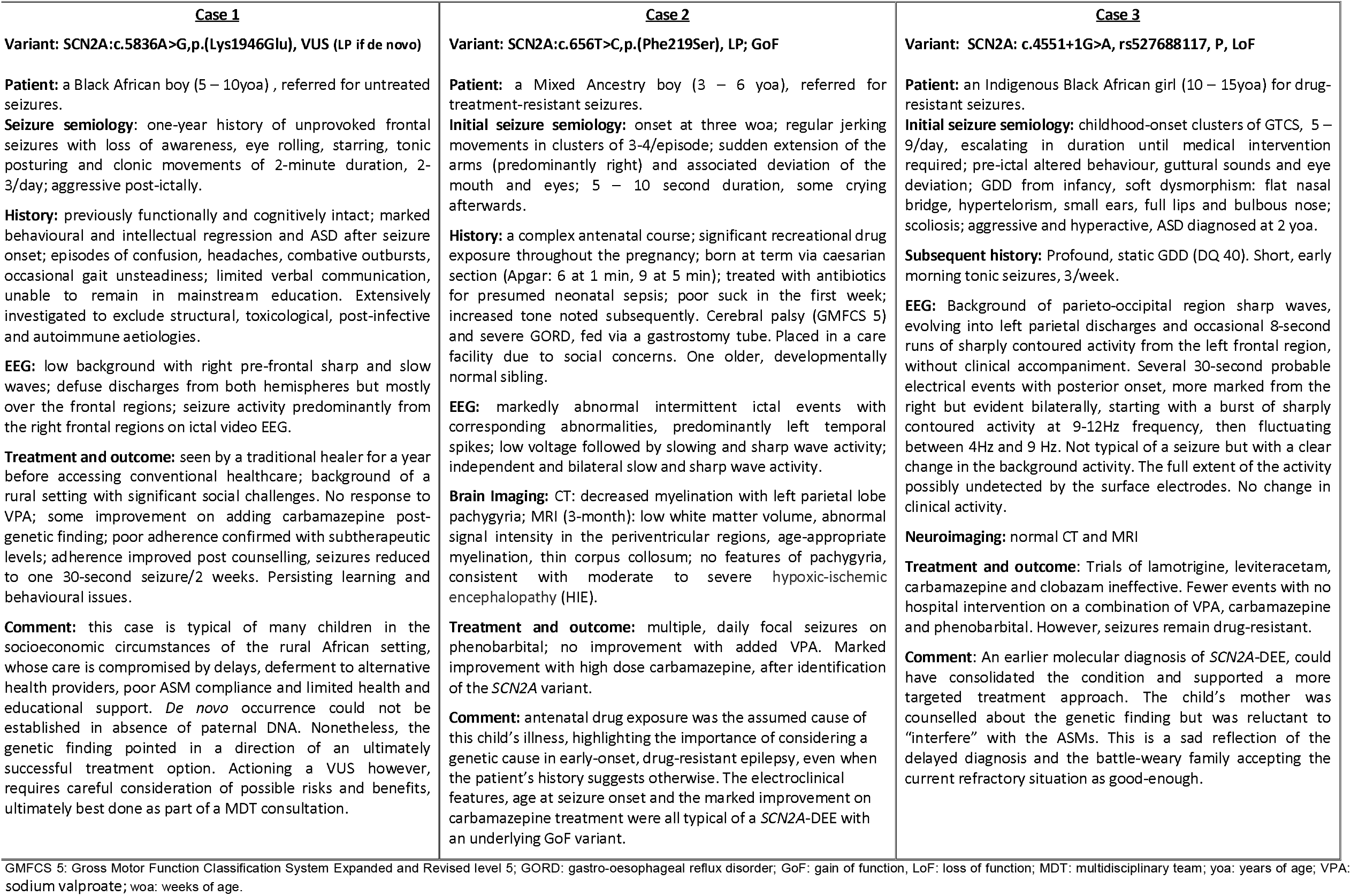
The “think-genetics” (TG) decision tree for recognition and genetic diagnosis of DEE in a low-income setting. *e.g., hand or arm stereotypies, gait abnormalities, eyelid myoclonia; ** panel of 32 genes with implication for precision treatment, and those commonly implicated in DEEs (Supplemental. Notes – 4); AD: attention difficulties, ASD: autism spectrum disorder, AOO: age of onset; CMA: chromosomal microarray, ES: exome sequencing, GDD: global developmental delay, GS: genome sequencing, ID/DD: intellectual disability/developmental delay; MDT: multidisciplinary team, VUS: variant of uncertain significance.

1. **Clinical features associated with the presence of P/LP variants in our study:** neonatal or infantile-onset, drug-resistant seizures are most likely to have a currently identifiable genetic cause(31). We identified movement abnormalities (stereotypical hand and arm movements, gait abnormalities, eyelid myoclonia), attention difficulties, ASD, and dysmorphic features as additional strong indicators of a genetic aetiology. Structural brain anomalies were excluded from this model, despite the strong statistical association with CNVs (Figure 3), as the description included non-specific features such as brain atrophy and thinning corpus, making it a relatively weak criterion for a genetic aetiology. Moreover, structural anomalies are identified by imaging and thus cannot be described as a simple diagnostic marker.
2. **Precision medicine implications:** initial testing with a “starter DEE panel” is more affordable and easier to set up locally than ES or GS. Almost all P/LP SNV/indels in our study were detected in 12 genes included in the NGS panel, listed among the 20 top-yielding DEE-associated genes, in published large-scale studies (9, 32, 33). Therefore, initial testing with a small panel of genes selected on the basis of clinical actionability and frequency of association, should solve a good proportion of the DEE cases, with possible options for precision treatment (Suppl. Note 4). This has been previously suggested by other investigators(34, 35).
3. **Choice of initial genetic testing modality:** we suggest CMA testing as the initial genetic investigation for patients with dysmorphic features and GDD prior to seizure onset, as these were prominent in patients with CNVs in our study. If negative, NGS-based testing for SNV/indels should follow. Conversely, patients with neonatal/infantile-onset seizures and normal prior development, should be tested with NGS first (starter panel) followed by CMA, if negative. MDT consultation should precede further testing (ES or GS), as additional information may have become available, re-directing the diagnostic focus away from a genetic cause.

## DISCUSSION

They findings of our study demonstrate first-hand that access to genetic testing for DEE, in the under-resourced, tertiary hospital setting in SA carries similar clinical and economic benefits to those described in the HICs (9, 36), justifying the cost of NGS and CMA.

The yield of P/LP variants in our study (17%) was lower than anticipated based on our panel size and published reports of similar testing in DEE, reporting a diagnostic yield of 20 – 45% (32, 35). This is likely due to the broad inclusion criteria and complexities of the local patient referral system. Our findings did, however, mirror international conclusions in terms of the diagnostic yield and age at seizure onset. Most of the P/LP variants in our study were identified in children with neonatal and infantile-onset seizures (50%) (Table 1)(31). The top four yielding genes in our panel (*SCN1A, STXPB1, SCN2A, KCNT1*) accounted for 50% of the solved cases (20/40) and 69% of the detected P/LP SNV/indels, also in keeping with the published DEE research(34, 37). Conspicuously absent from our group were pathogenic variants in *KCNQ2* which ranks among the five top-yielding genes in large-scale studies, and is linked to a phenotypic spectrum spanning the benign familial and the severe *de novo* DEE phenotypes (9, 32, 33). The reasons for this bear further investigation but may be related to our relatively small cohort size and possible patient exclusion based on presumption of infection or hypoxia, as the most common local aetiologies of neonatal seizures.

Apart from the *NARS* variant detected with ES, all the P/LP SNV/indels in our study were identified on panel testing, emphasising its utility. Gene panels are less expensive than ES or GS, require less skill and computational power to analyse, yield fewer VUS and almost no incidental findings. This is relevant in the African setting, as there is limited, publicly accessible data on allele frequencies in African populations, especially in SSA. D*e novo* occurrence, as the major criterion for variant interpretation in DEEs(38, 39) is often difficult to establish in the African setting, with its high prevalence of the so-called “orphan households”(5, 40). Therefore, we propose a small panel of 32 DEE genes, as a pragmatic and high-yielding first-tier genetic test for DEEs in LMICs. If negative, this would be followed by ES (if accessible), if there is still an indication and budget for further testing (Figure 4).

The proportion of the detected P/LP CNVs was high (12 out of 78 CMA-tested patients, i.e., 15%), mostly diagnostic of classical microdeletion/duplication syndromes (1p36, 22q11dup. Suppl. Tables 1 and 2). In a well-resourced setting, these patients would have been tested with an early diagnostic CMA and excluded from the study. Our patients, however, were referred to the Epilepsy Service for seizures as the initial presentation or major concern, and their care focused on the acute needs such as seizure control, development, respiratory or feeding problems. We ascribe the late diagnoses to the subtle presentation or incomplete penetrance in some cases. We did not exclude these patients from subsequent analyses, as our goal was to assess a real-life patient population in a paediatric epilepsy clinic in SA. The CNV detection highlighted the need to review the current local referral and clinical re-assessment protocols, incorporating CMA testing for epilepsy-plus phenotypes, especially those with ID and dysmorphism (41).

Of the 41 children with P/LP findings, 26 had variants with treatment implications (Suppl. Tables 1 and 2). The growing understanding of the mechanisms of variant pathogenicity and effect on ASMs, with treatment recommendations or contraindication based on the underlying genotype, is perhaps the most compelling rationale for availability of clinical genetic testing, especially for the DEEs (9, 42). The relevance of the functional effect of the underlying variant to the choice of treatment was demonstrated in our study by three patients *SCN2A* variants, each with a different variant type, age of seizure onset and response to treatment (Figure 1, Suppl. Table 1)(9, 11). These cases are an exciting example of precision treatment, often considered beyond the reach of healthcare in Africa.

Compared to similarly presenting patients in the HICs, the children in our study obtained their genetic diagnoses late, years after the onset of epilepsy (Table 1). The median age at molecular diagnosis among the infantile-onset group was 108 months (IQR: 96,156) compared to the median age of 9 months (IQR: 3,17), at seizure onset. However, even the late genetic diagnosis led to treatment adjustments and positive changes in the lives of the patients and the families. Even in absence of direct implications for treatment, the value of the diagnostic closure is vastly underestimated, especially in resource-constrained settings where genetic testing may be viewed as non-essential. Anecdotal feedback from the recruiting clinicians conveyed the profound relief and gratitude from the parents, for whom knowing the reason behind their child’s illness brought closure and focus on the way forward. The clinicians too, were grateful to have the answers and excited to gain new and useful knowledge. Few - if any – other research studies in that environment to date have elicited an equally positive response and ongoing enquiry from the families and the clinicians.

One may question the economic feasibility of weighing down the hospital budgets in resource-constrained settings with an expensive genetic investigation, which carries a realistic pick-up rate of less than 40%, even in a well-phenotyped and appropriately referred patient population. However, the overall expenses incurred during the so-called diagnostic odyssey are exceedingly high and the downstream savings carried by a molecular diagnosis outweigh the cost of testing(43, 44). At the time of writing this manuscript, the combined cost of a gene panel and a CMA in SA amounted to approximately 20,000ZAR (∼1,300USD). This is similar to the cost of a single brain MRI, often ordered more than once during the course of the child’s illness, as a standard investigation for DEE. Added to this are the costly metabolic screens (∼200USD per test) and hospital admissions.

In the SA daily clinical practice, recognition of possible signs of a genetic epilepsy is complicated by the layering effects of TB, HIV, parasitic and other febrile illness, perinatal insult, as well as poor nutrition and other complications of the socio-economic circumstance. Based on our findings, we designed the simple TG decision tree, which could guide the primary care clinician in Africa in recognising signs of a possible genetic aetiology, amongst the prevalence of seizures with an acquired cause. These patients could then be triaged correctly and much earlier on, for specialist consultation, correct intervention and, ideally, genetic testing. The few analysable variables and relatively small cohort size limited our ability to draw decisive conclusions from the multivariate analysis in this study. Nonetheless, our findings suggest that in addition to neonatal- or infantile-onset drug-resistant seizures, movement abnormalities, attention difficulties, ASD, dysmorphic features and GDD are strong additional indicators of a genetic aetiology, regardless of age at seizure-onset (Figure 4).

## CONCLUSION

To our knowledge, this is the first published study of the genetic underpinnings of DEE patients in SSA. An important characteristic of our study group, compared to the investigations conducted in well-resourced settings, was the absence of selection bias imposed by prior genetic testing. The convoluted diagnostic history of many children in our study, reflects the time delays between seizure onset, seeking medical help and eventual correct diagnosis and treatment. Our TG decision tree may help to limit such delays, by simplifying decision-making and setting the clinician at patient entry on a correct diagnostic course from as early as the first visit.

## Data Availability

Tables listing deidentified clinical information, genetic variant details, statistical summaries and regression analyses are included in the Supplemental Information. Deidentified raw data may be available on request from the corresponding author.

## STUDY APPROVAL

The study was approved by the Human Research Ethics Committee of the University of Cape Town (HREC REF: 232/2015). Written informed consent (and assent, were appropriate) was obtained from probands and parents, prior to participation in the study.

## AUTHOR CONTRIBUTIONS

Conceptualization: A.I.E, H.C.M., R.R., J.M.W., G.L.C.; Data curation: A.I.E., G.R., M.W., J.M.W.; Formal analysis: N.T., M.C.A., J.D.C., J.G., M.J.B., D.A.N.; Funding acquisition: R.R., J.M.W., G.L.C.; Investigation: A.I.E., M.W., G.R., M.C.A., E.E.A., A.R., J.L.W., G.L.C.; Resources: H.M.C., J.M.W., G.L.C.; Software: R.J.B., J.D.C., J.G.; Supervision: R.R., J.M.W, G.L.C.; Visualization: A.I.E., R.J.B., G.L.C.; Writing-original draft: A.I.E.; Writing-review & editing: A.I.E., N.T., R.J.B., H.C.M., R.R., J.M.W., G.L.C.

## Acknowledgements

We thank all the children and parents for participation and keen interest in the study, which made obtaining and communicating meaningful results so gratifying. We are also grateful to the staff in the Epilepsy Clinic at the Red Cross Children’s War Memorial Hospital in Cape Town for the effort put into patient recruitment, obtaining informed consent and browsing patient file archives, adding to their already heavy workload.

Exome sequencing was provided by the University of Washington Center for Mendelian Genomics (UW-CMG) and was funded by NHGRI and NHLBI grants UM1 HG006493 and U24 HG008956. The content is solely the responsibility of the authors and does not necessarily represent the official views of the National Institutes of Health.

