## Supplementary material for "Precision medicine for developmental and epileptic encephalopathies in Africa – strategies for a resource-limited setting"

**SUPPLEMENTAL INFORMATION**

**CONTENT: Page**

**Table 1.** Clinical Information for patients with identified P/LP variants and VUS of interest **- separate spreadsheet**

**Table 2**. Genetic Variants detected with the gene panel, ES and CMA (P/LP and VUS) **- separate spreadsheet**

**Table 3**. Summary Statistics Comparing the Clinical Characteristics of Patients with Candidate

SNVs/indels and Patients with No Detected Candidate SNVs/indels. **2**

**Table 4.** Summary Statistics Comparing the Clinical Characteristics of Patients with Candidate

CNVs and Patients with no Detected Candidate CNVs. **4**

**Table 5.** Summary Statistics Comparing the Clinical Characteristics of Patients with Candidate Variants

(SNVs/indels and CNVs) and Patients with No Detected Candidate Variants. **6**

**Table 6**. Multiple Linear Regression Modelling to Assess Associations Between Selected Clinical

Features and a Detected Candidate SNV/indel. **8**

**Table 7.** Multiple Linear Regression Modelling to Assess Associations Between Selected Clinical

Features and a Detected Candidate CNV. **8**

**Table 8.** Multiple Linear Regression Modelling to Assess Associations Between Selected Clinical

Features and the Detected Candidate Genetic Variant (SNV/indel or CNV). **8**

**Figure 1**. Seizure types in P/LP variant-positive (A) and P/LP variant-negative (B) children,

stratified per age of seizure onset: neonatal, infantile and childhood	**9**

**Figure 2**: Multiple Linear Regression Modelling to Assess Associations Between Selected Clinical

Features and the Detected Candidate Genetic Variant (SNV/indel or CNV). **9**

**Supplementary Notes:**

**Supplementary Note 1:** Essential Clinical Info Requested	**10**

**Supplementary Note 2:** DEE MIP panel (71 genes)	**11**

**Supplementary Note 3:** DEE-related genes with high coverage on the custom array	**11**

**Supplementary Note 4:** DEE starter panel design	**11**

**Table 3.** Summary Statistics Comparing the Clinical Characteristics of Patients with Candidate SNVs/indels and Patients with No Detected Candidate SNVs/indels.

|  | **All patients**  N = 235*^1^* | **Patients with no candidate SNVs/indels**  N = 195*^1^* | **Patients with candidate SNVs/indels**  N = 40*^1^* | **p-value***^2^* |
| --- | --- | --- | --- | --- |
| seizure type at onset: febrile | 39 (17%) | 30 (15%) | 9 (22%) | 0.3 |
| seizure type at onset: focal | 57 (24%) | 40 (21%) | 17 (42%) | 0.003 |
| seizure type at onset: hemiclonic | 3 (1.3%) | 1 (0.5%) | 2 (5.0%) | 0.076 |
| seizure type at onset: spasms | 33 (14%) | 29 (15%) | 4 (10%) | 0.4 |
| seizure type at onset: other | 21 (8.9%) | 18 (9.2%) | 3 (7.5%) | >0.9 |
| seizure type at onset: generalised | 99 (42%) | 77 (39%) | 22 (55%) | 0.070 |
| seizure type at onset: tonic | 20 (8.5%) | 17 (8.7%) | 3 (7.5%) | >0.9 |
| seizure type at onset: generalised clonic | 4 (1.7%) | 2 (1.0%) | 2 (5.0%) | 0.14 |
| seizure type at onset: generalised tonic clonic | 71 (30%) | 54 (28%) | 17 (42%) | 0.063 |
| seizure type at onset: generalised myoclonic | 19 (8.1%) | 15 (7.7%) | 4 (10%) | 0.5 |
| seizure type at onset: generalised absence | 2 (0.9%) | 1 (0.5%) | 1 (2.5%) | 0.3 |
| seizure type at onset: generalised atonic | 7 (3.0%) | 7 (3.6%) | 0 (0%) | 0.6 |
| current seizure type: febrile | 17 (7.2%) | 13 (6.7%) | 4 (10%) | 0.5 |
| current seizure type: focal | 64 (27%) | 45 (23%) | 19 (48%) | 0.002 |
| current seizure type: hemiclonic | 3 (1.3%) | 0 (0%) | 3 (7.5%) | 0.005 |
| current seizure type: spasms | 7 (3.0%) | 6 (3.1%) | 1 (2.5%) | >0.9 |
| current seizure type: other | 11 (4.7%) | 7 (3.6%) | 4 (10%) | 0.10 |
| current seizure type: generalised | 117 (50%) | 92 (47%) | 25 (62%) | 0.078 |
| current seizure type: generalised tonic | 42 (18%) | 34 (17%) | 8 (20%) | 0.7 |
| current seizure type: generalised clonic | 6 (2.6%) | 4 (2.1%) | 2 (5.0%) | 0.3 |
| current seizure type: generalised tonic clonic | 84 (36%) | 69 (35%) | 15 (38%) | 0.8 |
| current seizure type: generalised myoclonic | 53 (23%) | 38 (19%) | 15 (38%) | 0.013 |
| current seizure type: generalised absence | 18 (7.7%) | 14 (7.2%) | 4 (10%) | 0.5 |
| current seizure type: generalised atonic | 22 (9.4%) | 17 (8.7%) | 5 (12%) | 0.5 |
| current seizure type: generalised atonic | 10 (4.3%) | 9 (4.6%) | 1 (2.5%) | >0.9 |
| seizure trigger | 82 (41%) | 62 (38%) | 20 (54%) | 0.065 |
| status epilepticus (SE) | 84 (41%) | 62 (37%) | 22 (58%) | 0.017 |
| developmental delay before seizure onset | 58 (29%) | 49 (30%) | 9 (23%) | 0.4 |
| gait disorder before seizure onset | 9 (4.3%) | 7 (4.0%) | 2 (5.3%) | 0.7 |
| impaired motor dev. before seizure onset | 35 (17%) | 32 (19%) | 3 (8.3%) | 0.12 |
| cognitive delay prior to seizure onset | 33 (17%) | 28 (18%) | 5 (14%) | 0.6 |
| neuro-regression | 83 (39%) | 64 (37%) | 19 (48%) | 0.2 |
| neuro-regression associated with poor seizure control / clustering | 60 (77%) | 46 (75%) | 14 (82%) | 0.7 |
| family history of seizures | 55 (26%) | 42 (24%) | 13 (33%) | 0.2 |
| family history of febrile seizures | 11 (5.3%) | 7 (4.1%) | 4 (11%) | 0.11 |
| family history of developmental problems | 7 (3.4%) | 6 (3.6%) | 1 (2.6%) | >0.9 |
| dysmorphic features | 26 (11%) | 19 (9.7%) | 7 (18%) | 0.2 |
| visual impairment | 29 (13%) | 23 (13%) | 6 (15%) | 0.8 |
| focal neurological deficit | 26 (12%) | 21 (12%) | 5 (13%) | 0.8 |
| ophthalmological abnormalities | 16 (7.4%) | 13 (7.4%) | 3 (7.7%) | >0.9 |
| normal EEG background | 105 (50%) | 81 (47%) | 24 (62%) | 0.10 |
| Evolving EEG background | 50 (46%) | 34 (43%) | 16 (55%) | 0.3 |
| interictal epileptiform activity | 96 (45%) | 81 (47%) | 15 (38%) | 0.4 |
| Hypsarrythmia | 18 (7.7%) | 17 (8.7%) | 1 (2.5%) | 0.3 |
| Burst-suppression | 22 (9.4%) | 20 (10%) | 2 (5.0%) | 0.4 |
| electro-decrements | 5 (2.1%) | 4 (2.1%) | 1 (2.5%) | >0.9 |
| modified hypsarrythmia | 9 (3.8%) | 8 (4.1%) | 1 (2.5%) | >0.9 |
| other EEG patterns | 13 (5.5%) | 12 (6.2%) | 1 (2.5%) | 0.7 |
| Structural brain anomalies | 12 (5.1%) | 8 (4.1%) | 4 (10%) | 0.13 |
| global atrophy | 18 (7.7%) | 13 (6.7%) | 5 (12%) | 0.2 |
| delayed myelination | 3 (1.3%) | 1 (0.5%) | 2 (5.0%) | 0.076 |
| thinning corpus callosum | 11 (4.7%) | 7 (3.6%) | 4 (10%) | 0.10 |
| focal cortical dysplasia | 5 (2.1%) | 5 (2.6%) | 0 (0%) | 0.6 |
| evidence of hypoxic-ischemic encephalopathy (HIE) | 17 (7.2%) | 11 (5.6%) | 6 (15%) | 0.048 |
| calcification | 3 (1.3%) | 3 (1.5%) | 0 (0%) | >0.9 |
| other findings on imaging | 58 (25%) | 50 (26%) | 8 (20%) | 0.5 |
| movement disorder: chorea | 2 (0.9%) | 1 (0.5%) | 1 (2.5%) | 0.3 |
| movement disorder: dystonia | 6 (2.6%) | 5 (2.6%) | 1 (2.5%) | >0.9 |
| movement disorder: stereotypy | 7 (3.0%) | 5 (2.6%) | 2 (5.0%) | 0.3 |
| movement disorder: tremor | 1 (0.4%) | 0 (0%) | 1 (2.5%) | 0.2 |
| movement disorder: other | 4 (1.7%) | 1 (0.5%) | 3 (7.5%) | 0.016 |
| crouched gait | 7 (3.3%) | 4 (2.3%) | 3 (7.7%) | 0.11 |
| sleep cycle disturbance | 14 (6.6%) | 9 (5.2%) | 5 (13%) | 0.14 |
| no psychiatric disorders | 130 (55%) | 110 (56%) | 20 (50%) | 0.5 |
| hyperactivity | 44 (19%) | 34 (17%) | 10 (25%) | 0.3 |
| depression | 1 (0.4%) | 0 (0%) | 1 (2.5%) | 0.2 |
| attention deficit | 21 (8.9%) | 13 (6.7%) | 8 (20%) | 0.013 |
| autism spectrum | 30 (13%) | 21 (11%) | 9 (22%) | 0.043 |
| other psychiatric disorders | 8 (3.4%) | 8 (4.1%) | 0 (0%) | 0.4 |
| behavioural problems | 61 (31%) | 46 (29%) | 15 (41%) | 0.2 |
| HIV exposure | 15 (6.7%) | 13 (7.1%) | 2 (5.0%) | 0.8 |
| Tuberculosis | 5 (2.2%) | 4 (2.2%) | 1 (2.6%) | >0.9 |
| previous traumatic brain injury | 4 (1.8%) | 3 (1.6%) | 1 (2.6%) | 0.5 |
| age at first seizure | 8 (3, 18) | 9 (4, 18) | 5 (2, 9) | 0.017 |
| *^1^*n (%); Median (IQR) *^2^*Pearson's Chi-squared test; Fisher's exact test; Wilcoxon rank sum test | | | | |

**Table 4**. Summary Statistics Comparing the Clinical Characteristics of Patients with Candidate CNVs and	Patients	with no Detected Candidate CNVs

|  | **All patients**  N = 235*^1^* | **Patients with no candidate CNVs**  N = 223*^1^* | **Patients with candidate CNVs**  N = 12*^1^* | **p-value***^2^* |
| --- | --- | --- | --- | --- |
| seizure type at onset: febrile | 39 (17%) | 38 (17%) | 1 (8.3%) | 0.7 |
| seizure type at onset: focal | 57 (24%) | 55 (25%) | 2 (17%) | 0.7 |
| seizure type at onset: hemiclonic | 3 (1.3%) | 3 (1.3%) | 0 (0%) | >0.9 |
| seizure type at onset: spasms | 33 (14%) | 32 (14%) | 1 (8.3%) | >0.9 |
| seizure type at onset: other | 21 (8.9%) | 19 (8.5%) | 2 (17%) | 0.3 |
| seizure type at onset: generalised | 99 (42%) | 95 (43%) | 4 (33%) | 0.5 |
| seizure type at onset: generalised tonic | 20 (8.5%) | 19 (8.5%) | 1 (8.3%) | >0.9 |
| seizure type at onset: generalised clonic | 4 (1.7%) | 4 (1.8%) | 0 (0%) | >0.9 |
| seizure type at onset: generalised tonic clonic | 71 (30%) | 68 (30%) | 3 (25%) | >0.9 |
| seizure type at onset: generalised myoclonic | 19 (8.1%) | 17 (7.6%) | 2 (17%) | 0.3 |
| seizure type at onset: generalised absence | 2 (0.9%) | 2 (0.9%) | 0 (0%) | >0.9 |
| seizure type at onset: generalised atonic | 7 (3.0%) | 7 (3.1%) | 0 (0%) | >0.9 |
| seizure type at onset: other | 4 (1.7%) | 4 (1.8%) | 0 (0%) | >0.9 |
| current seizure type: febrile | 17 (7.2%) | 16 (7.2%) | 1 (8.3%) | 0.6 |
| current seizure type: focal | 64 (27%) | 63 (28%) | 1 (8.3%) | 0.2 |
| current seizure type: hemiclonic | 3 (1.3%) | 3 (1.3%) | 0 (0%) | >0.9 |
| current seizure type: spasms | 7 (3.0%) | 7 (3.1%) | 0 (0%) | >0.9 |
| current seizure type: other | 11 (4.7%) | 10 (4.5%) | 1 (8.3%) | 0.4 |
| current seizure type: generalised | 117 (50%) | 109 (49%) | 8 (67%) | 0.2 |
| current seizure type: tonic | 42 (18%) | 41 (18%) | 1 (8.3%) | 0.7 |
| current seizure type: generalised clonic | 6 (2.6%) | 6 (2.7%) | 0 (0%) | >0.9 |
| current seizure type: generalised tonic clonic | 84 (36%) | 80 (36%) | 4 (33%) | >0.9 |
| current seizure type: generalised myoclonic | 53 (23%) | 49 (22%) | 4 (33%) | 0.5 |
| current seizure type: generalised absence | 18 (7.7%) | 18 (8.1%) | 0 (0%) | 0.6 |
| current seizure type: generalised atonic | 22 (9.4%) | 20 (9.0%) | 2 (17%) | 0.3 |
| current seizure type: generalised atonic | 10 (4.3%) | 10 (4.5%) | 0 (0%) | >0.9 |
| seizure trigger | 82 (41%) | 76 (40%) | 6 (55%) | 0.4 |
| status epilepticus (SE) | 84 (41%) | 82 (42%) | 2 (20%) | 0.2 |
| Developmental delay before seizure onset | 58 (29%) | 52 (27%) | 6 (60%) | 0.034 |
| gait disorder before seizure onset | 9 (4.3%) | 9 (4.5%) | 0 (0%) | >0.9 |
| impaired motor development before seizure onset | 35 (17%) | 29 (15%) | 6 (60%) | 0.002 |
| cognitive delay prior to seizure onset | 33 (17%) | 27 (15%) | 6 (60%) | 0.002 |
| neuro-regression | 83 (39%) | 80 (39%) | 3 (27%) | 0.5 |
| neuro-regression associated with poor seizure control / clustering | 60 (77%) | 58 (77%) | 2 (67%) | 0.6 |
| family history of seizures | 55 (26%) | 53 (26%) | 2 (20%) | >0.9 |
| family history of febrile seizures | 11 (5.3%) | 10 (5.0%) | 1 (10%) | 0.4 |
| family history of developmental problems | 7 (3.4%) | 7 (3.6%) | 0 (0%) | >0.9 |
| dysmorphic features | 26 (11%) | 20 (9.0%) | 6 (50%) | <0.001 |
| visual impairment | 29 (13%) | 26 (13%) | 3 (27%) | 0.2 |
| movement abnormality | 35 (16%) | 32 (15%) | 3 (25%) | 0.4 |
| focal neurological deficit | 26 (12%) | 24 (12%) | 2 (18%) | 0.6 |
| ophthalmological abnormalities | 16 (7.4%) | 13 (6.3%) | 3 (30%) | 0.030 |
| normal EEG background | 105 (50%) | 99 (49%) | 6 (60%) | 0.5 |
| Evolving EEG background | 50 (46%) | 48 (46%) | 2 (50%) | >0.9 |
| interictal epileptiform activity | 96 (45%) | 93 (46%) | 3 (27%) | 0.4 |
| Was the ictal activity recorded? | 62 (30%) | 60 (31%) | 2 (20%) | 0.7 |
| no specific EEG pattern | 158 (67%) | 150 (67%) | 8 (67%) | >0.9 |
| Hypsarrythmia | 18 (7.7%) | 16 (7.2%) | 2 (17%) | 0.2 |
| Burst-suppression | 22 (9.4%) | 21 (9.4%) | 1 (8.3%) | >0.9 |
| electro-decrements | 5 (2.1%) | 5 (2.2%) | 0 (0%) | >0.9 |
| modified hypsarrythmia | 9 (3.8%) | 9 (4.0%) | 0 (0%) | >0.9 |
| other EEG patterns | 13 (5.5%) | 13 (5.8%) | 0 (0%) | >0.9 |
| brain abnormalities on imaging | 89 (44%) | 83 (44%) | 6 (50%) | 0.8 |
| global atrophy | 18 (7.7%) | 16 (7.2%) | 2 (17%) | 0.2 |
| delayed myelination | 3 (1.3%) | 2 (0.9%) | 1 (8.3%) | 0.15 |
| thinning corpus callosum | 11 (4.7%) | 7 (3.1%) | 4 (33%) | 0.001 |
| focal cortical dysplasia | 5 (2.1%) | 5 (2.2%) | 0 (0%) | >0.9 |
| evidence of hypoxic-ischemic encephalopathy (HIE) | 17 (7.2%) | 16 (7.2%) | 1 (8.3%) | 0.6 |
| calcification | 3 (1.3%) | 3 (1.3%) | 0 (0%) | >0.9 |
| movement disorder | 20 (9.2%) | 17 (8.3%) | 3 (27%) | 0.069 |
| movement disorder: chorea | 2 (0.9%) | 1 (0.4%) | 1 (8.3%) | 0.10 |
| movement disorder: dystonia | 6 (2.6%) | 4 (1.8%) | 2 (17%) | 0.032 |
| movement disorder: stereotypy | 7 (3.0%) | 6 (2.7%) | 1 (8.3%) | 0.3 |
| movement disorder: tremor | 1 (0.4%) | 1 (0.4%) | 0 (0%) | >0.9 |
| crouched gait | 7 (3.3%) | 7 (3.4%) | 0 (0%) | >0.9 |
| sleep cycle disturbance | 14 (6.6%) | 13 (6.4%) | 1 (10%) | 0.5 |
| no psychiatric disorders | 130 (55%) | 123 (55%) | 7 (58%) | 0.8 |
| hyperactivity | 44 (19%) | 42 (19%) | 2 (17%) | >0.9 |
| depression | 1 (0.4%) | 1 (0.4%) | 0 (0%) | >0.9 |
| attention deficit | 21 (8.9%) | 21 (9.4%) | 0 (0%) | 0.6 |
| autism spectrum | 30 (13%) | 28 (13%) | 2 (17%) | 0.7 |
| behavioural problems | 61 (31%) | 58 (31%) | 3 (30%) | >0.9 |
| HIV exposure | 15 (6.7%) | 15 (7.1%) | 0 (0%) | >0.9 |
| Tuberculosis | 5 (2.2%) | 5 (2.3%) | 0 (0%) | >0.9 |
| previous traumatic brain injury | 4 (1.8%) | 3 (1.4%) | 1 (8.3%) | 0.2 |
| age at first seizure | 8 (3, 18) | 8 (3, 18) | 7 (6, 18) | 0.7 |
| *^1^*n (%); Median (IQR) *^2^*Pearson's Chi-squared test; Fisher's exact test; Wilcoxon rank sum test | | | | |

**Table 5**. Summary Statistics Comparing the Clinical Characteristics of Patients with Candidate Variants

(SNVs/indels and CNVs) and Patients with No Detected Candidate Variants.

|  | **All patients**  N = 235*^1^* | **Patients with no candidate SNVs/indels or CNVs**  N = 183*^1^* | **Patients with candidate**  **SNVs/indels and CNVs** N = 52*^1^* | **p-value***^2^* |
| --- | --- | --- | --- | --- |
| seizure type at onset: febrile | 39 (17%) | 29 (16%) | 10 (19%) | 0.6 |
| seizure type at onset: focal | 57 (24%) | 38 (21%) | 19 (37%) | 0.019 |
| seizure type at onset: hemiclonic | 3 (1.3%) | 1 (0.5%) | 2 (3.8%) | 0.12 |
| seizure type at onset: spasms | 33 (14%) | 28 (15%) | 5 (9.6%) | 0.3 |
| seizure type at onset: other | 21 (8.9%) | 16 (8.7%) | 5 (9.6%) | 0.8 |
| seizure type at onset: generalised | 99 (42%) | 73 (40%) | 26 (50%) | 0.2 |
| seizure type at onset: generalised tonic | 20 (8.5%) | 16 (8.7%) | 4 (7.7%) | >0.9 |
| seizure type at onset: generalised clonic | 4 (1.7%) | 2 (1.1%) | 2 (3.8%) | 0.2 |
| seizure type at onset: generalised tonic clonic | 71 (30%) | 51 (28%) | 20 (38%) | 0.14 |
| seizure type at onset: generalised myoclonic | 19 (8.1%) | 13 (7.1%) | 6 (12%) | 0.4 |
| seizure type at onset: generalised absence | 2 (0.9%) | 1 (0.5%) | 1 (1.9%) | 0.4 |
| seizure type at onset: generalised atonic | 7 (3.0%) | 7 (3.8%) | 0 (0%) | 0.4 |
| seizure type at onset: other | 4 (1.7%) | 3 (1.6%) | 1 (1.9%) | >0.9 |
| current seizure type: febrile | 17 (7.2%) | 12 (6.6%) | 5 (9.6%) | 0.5 |
| current seizure type: focal | 64 (27%) | 44 (24%) | 20 (38%) | 0.039 |
| current seizure type: hemiclonic | 3 (1.3%) | 0 (0%) | 3 (5.8%) | 0.010 |
| current seizure type: spasms | 7 (3.0%) | 6 (3.3%) | 1 (1.9%) | >0.9 |
| current seizure type: generalised | 117 (50%) | 84 (46%) | 33 (63%) | 0.025 |
| current seizure type: generalised tonic | 42 (18%) | 33 (18%) | 9 (17%) | >0.9 |
| current seizure type: generalised clonic | 6 (2.6%) | 4 (2.2%) | 2 (3.8%) | 0.6 |
| current seizure type: generalised tonic_clonic | 84 (36%) | 65 (36%) | 19 (37%) | 0.9 |
| current seizure type: generalised myoclonic | 53 (23%) | 34 (19%) | 19 (37%) | 0.006 |
| current seizure type: generalised absence | 18 (7.7%) | 14 (7.7%) | 4 (7.7%) | >0.9 |
| current seizure type: generalised atonic | 22 (9.4%) | 15 (8.2%) | 7 (13%) | 0.3 |
| current seizure type: generalised atonic | 10 (4.3%) | 9 (4.9%) | 1 (1.9%) | 0.7 |
| seizure trigger | 82 (41%) | 56 (36%) | 26 (54%) | 0.028 |
| status epilepticus (SE) | 84 (41%) | 60 (38%) | 24 (50%) | 0.14 |
| Developmental delay before seizure onset | 58 (29%) | 43 (28%) | 15 (31%) | 0.7 |
| gait disorder before seizure onset | 9 (4.3%) | 7 (4.3%) | 2 (4.2%) | >0.9 |
| impaired motor development before seizure onset | 35 (17%) | 26 (16%) | 9 (20%) | 0.6 |
| cognitive delay prior to seizure onset | 33 (17%) | 22 (15%) | 11 (24%) | 0.14 |
| neuro-regression | 83 (39%) | 61 (37%) | 22 (43%) | 0.5 |
| neuro-regression associated with poor seizure control / clustering | 60 (77%) | 44 (76%) | 16 (80%) | >0.9 |
| family history of seizures | 55 (26%) | 40 (24%) | 15 (31%) | 0.4 |
| family history of febrile seizures | 11 (5.3%) | 6 (3.7%) | 5 (11%) | 0.073 |
| family history of developmental problems | 7 (3.4%) | 6 (3.8%) | 1 (2.1%) | >0.9 |
| dysmorphic features | 26 (11%) | 13 (7.1%) | 13 (25%) | <0.001 |
| visual impairment | 29 (13%) | 20 (12%) | 9 (18%) | 0.3 |
| focal neurological deficit | 26 (12%) | 19 (11%) | 7 (14%) | 0.6 |
| ophthalmological abnormalities | 16 (7.4%) | 10 (6.0%) | 6 (12%) | 0.2 |
| normal EEG background | 105 (50%) | 75 (46%) | 30 (61%) | 0.062 |
| Evolving EEG background | 50 (46%) | 32 (43%) | 18 (55%) | 0.3 |
| interictal epileptiform activity | 96 (45%) | 78 (48%) | 18 (36%) | 0.14 |
| Was the ictal activity recorded? | 62 (30%) | 45 (28%) | 17 (35%) | 0.4 |
| no specific EEG pattern | 158 (67%) | 119 (65%) | 39 (75%) | 0.2 |
| Hypsarrythmia | 18 (7.7%) | 15 (8.2%) | 3 (5.8%) | 0.8 |
| Burst-suppression | 22 (9.4%) | 19 (10%) | 3 (5.8%) | 0.4 |
| electro-decrements | 5 (2.1%) | 4 (2.2%) | 1 (1.9%) | >0.9 |
| modified hypsarrythmia | 9 (3.8%) | 8 (4.4%) | 1 (1.9%) | 0.7 |
| Structural brain abnormalities on imaging | 12 (5.1%) | 3 (1.6%) | 9 (17%) | <0.001 |
| global atrophy | 18 (7.7%) | 11 (6.0%) | 7 (13%) | 0.083 |
| delayed myelination | 3 (1.3%) | 0 (0%) | 3 (5.8%) | 0.010 |
| thinning corpus callosum | 11 (4.7%) | 3 (1.6%) | 8 (15%) | <0.001 |
| focal cortical dysplasia | 5 (2.1%) | 5 (2.7%) | 0 (0%) | 0.6 |
| evidence of hypoxic-ischemic encephalopathy (HIE) | 17 (7.2%) | 10 (5.5%) | 7 (13%) | 0.066 |
| calcification | 3 (1.3%) | 3 (1.6%) | 0 (0%) | >0.9 |
| other findings on imaging | 58 (25%) | 45 (25%) | 13 (25%) | >0.9 |
| movement disorder | 39 (17%) | 17 (9.3%) | 22 (42%) | <0.001 |
| movement disorder: chorea | 2 (0.9%) | 0 (0%) | 2 (3.8%) | 0.048 |
| movement disorder: dystonia | 6 (2.6%) | 3 (1.6%) | 3 (5.8%) | 0.12 |
| movement disorder: stereotypy | 7 (3.0%) | 4 (2.2%) | 3 (5.8%) | 0.2 |
| movement disorder: tremor | 1 (0.4%) | 0 (0%) | 1 (1.9%) | 0.2 |
| crouched gait | 7 (3.3%) | 4 (2.4%) | 3 (6.1%) | 0.2 |
| sleep cycle disturbance | 14 (6.6%) | 8 (4.9%) | 6 (12%) | 0.092 |
| hyperactivity | 44 (19%) | 32 (17%) | 12 (23%) | 0.4 |
| depression | 1 (0.4%) | 0 (0%) | 1 (1.9%) | 0.2 |
| attention deficit | 21 (8.9%) | 13 (7.1%) | 8 (15%) | 0.094 |
| autism spectrum | 30 (13%) | 19 (10%) | 11 (21%) | 0.040 |
| behavioural problems | 61 (31%) | 43 (28%) | 18 (38%) | 0.2 |
| HIV exposure | 15 (6.7%) | 13 (7.6%) | 2 (3.8%) | 0.6 |
| Tuberculosis | 5 (2.2%) | 4 (2.3%) | 1 (2.0%) | >0.9 |
| previous traumatic brain injury | 4 (1.8%) | 2 (1.2%) | 2 (3.9%) | 0.2 |
| age at first seizure | 8 (3, 18) | 9 (4, 18) | 6 (3, 10) | 0.045 |
| *^1^*n (%); Median (IQR) *^2^*Pearson's Chi-squared test; Fisher's exact test; Wilcoxon rank sum test | | | | |

**Table 6**. Multiple Linear Regression Modelling to Assess Associations Between Selected Clinical Features and

a Detected Candidate SNV/indel.

| **Characteristic** | **OR***^1^* | **95% CI***^1^* | **p-value** |
| --- | --- | --- | --- |
| Autism spectrum | 2.24 | 0.74, 6.46 | 0.14 |
| Age at 1st Seizure | 0.98 | 0.95, 1.01 | 0.2 |
| Structural Brain Anomaly | 1.82 | 0.36, 8.28 | 0.4 |
| Movement Abnormality | 6.08 | 2.49, 15.2 | <0.001 |
| GDD | 0.41 | 0.13, 1.12 | 0.10 |
| Dysmorphism | 1.89 | 0.53, 6.30 | 0.3 |
| Attention Deficit | 3.96 | 1.24, 12.3 | 0.018 |
| *^1^*OR = Odds Ratio, CI = Confidence Interval | | | |

**Table 7.** Multiple Linear Regression Modelling to Assess Associations Between Selected Clinical Features and

a Detected Candidate CNV.

| **Characteristic** | **OR***^1^* | **95% CI***^1^* | **p-value** |
| --- | --- | --- | --- |
| Autism spectrum | 0.57 | 0.02, 4.92 | 0.7 |
| Age at 1st Seizure | 1.01 | 0.96, 1.05 | 0.5 |
| Structural Brain Anomaly | 22.8 | 3.25, 181 | 0.002 |
| Movement Abnormality | 0.52 | 0.05, 3.27 | 0.5 |
| GDD | 1.70 | 0.26, 9.91 | 0.6 |
| Dysmorphism | 4.52 | 0.63, 29.1 | 0.11 |
| *^1^*OR = Odds Ratio, CI = Confidence Interval | | | |

**Table 8.** Multiple Linear Regression Modelling to Assess Associations Between Selected Clinical Features and

a Detected Candidate Genetic Variant (SNV/indel or CNV).

| **Characteristic** | **OR***^1^* | **95% CI***^1^* | **p-value** |
| --- | --- | --- | --- |
| Autism spectrum | 2.11 | 0.72, 5.88 | 0.2 |
| Age at 1st Seizure | 0.99 | 0.96, 1.01 | 0.3 |
| Structural Brain Anomaly | 10.6 | 2.14, 78.4 | 0.007 |
| Movement Abnormality | 5.00 | 2.12, 11.9 | <0.001 |
| GDD | 0.57 | 0.21, 1.43 | 0.3 |
| Dysmorphism | 3.13 | 0.98, 9.90 | 0.050 |
| Attention Deficit | 2.83 | 0.93, 8.32 | 0.060 |
| *^1^*OR = Odds Ratio, CI = Confidence Interval | | | |

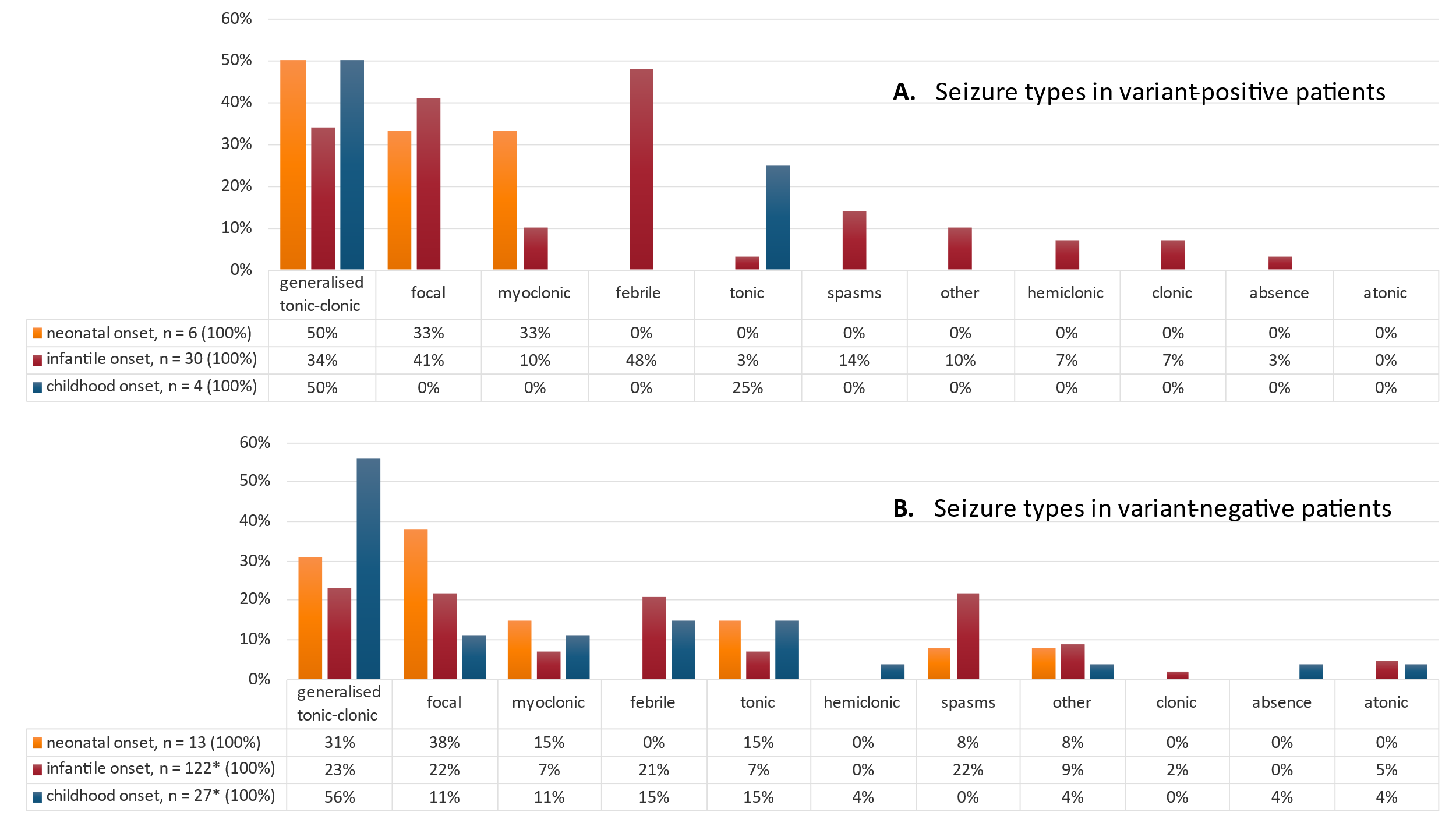

Figure 1. **Seizure types in P/LP variant-positive (A) and P/LP variant-negative (B) children, stratified per**

**age of seizure onset: neonatal, infantile and childhood.** *children with VUS were excluded.

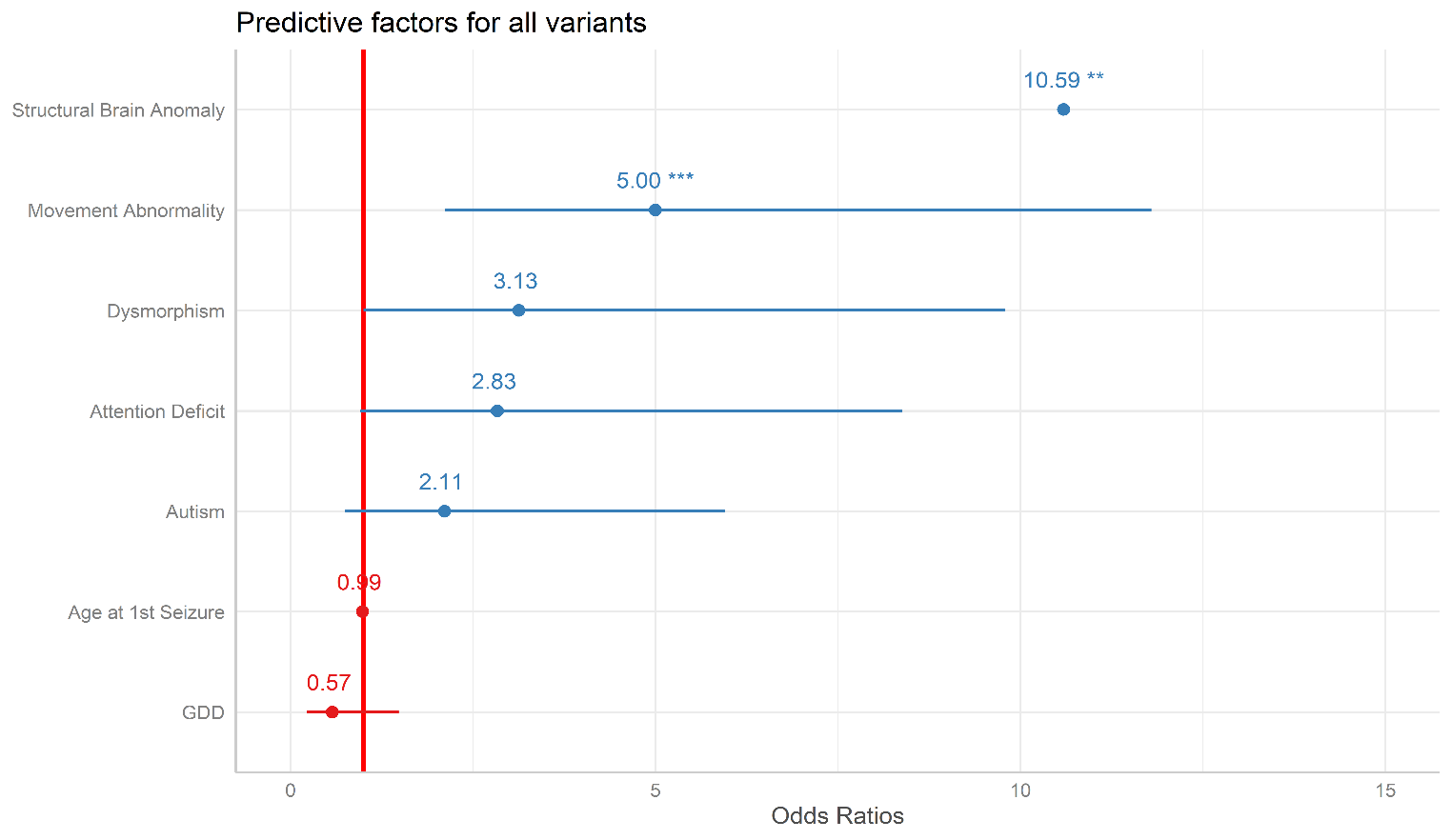

Figure 2: **Multiple Linear Regression modelling to assess associations between selected clinical features and presence of an identified SNV/indel or CNV (P/LP and VUS).**

**Supplementary Notes**

**Supplementary Note 1:** Essential Clinical Info Requested

DOB

Ethnicity

Neonatal complications

Age at 1st seizure

Age first seen

Initial seizure/s (type, frequency, special features e.g., hand movements)

Seizure evolution (type, frequency, special features e.g., hand movements)

Status epilepticus (number of events/duration)

Seizure triggers

ASMs trialled

Current ASMs

Ketogenic diet (trialled/ongoing/effect)

Response to treatment

History of non-adherence?

Development before seizure onset

Co-existing clinical features/conditions

Subsequent development

Seizure-associated neuroregression

Gait/movement abnormality

Other clinical features/conditions noted post seizure onset

Family history of seizures/epilepsy (details, if available)

EEG

Imaging: (type and findings)

Initial working diagnosis

Diagnostic genetic investigations and results

Final/current diagnosis

**Supplementary Note 2:** DEE MIP panel (71 genes):

Targeted NGS panel of 71 DEE-associated genes was performed using the previously described single molecule Molecular Inversion Probe (smMIP) technology(19). All exons and intron-exon boundaries (5-bp flanking sequences) were sequenced at 98% capture and 40X minimum depth of coverage (RefSeq, hg19 build)(20). NGS quality assessment, read alignment, depth of coverage, variant identification, annotation, prioritisation, and filtering was also performed using previously published methods(20–22). The VCF files were then subjected to further manual variant filtering and prioritisation.

*ALG13, ARX, ASH1L, ATP1A2, CACNA1A, CASK, CDKL5, CHD2, CLCN4, CUX2, DCX, DEPDC5, DMN1, DYRK1A, EEF1A2, FBXO28, FOXG1, FOXP1, GABRA1, GABRB1, GABRB2, GABRB3, GABRG2, GNAO1, GNB1, GRIN1, GRIN2A, GRIN2B, GRIN2D, HCN1, HCN2, HNRNPU, KCNA2, KCNB1, KCNH5, KCNQ2, KCNQ3. KCNT1, KIAA2022, MBD5, MECP2, MEF2C, NR2F1, PCDH19, PNKP, PNPO, PURA, RBFOX1, RORB, SCN1A, SCN1B, SCN2A, SCN3A, SCN8A, SIK1, SLC13A5, SLC1A2, SLC2A1, SLC35A2, SLC6A1, SMC1A, SNAP25, STX1B, STXBP1, SYN1, SYNGAP1, TBC1D24, TBL1XR1, TCF4, UBE3A, WDR45.*

**Supplementary Note 3: DEE-related genes with high coverage on the custom array (n=89 genes)**

This targeted custom array incorporated high-coverage tiling over epilepsy-associated genes (n=89), with one probe/150bp across the gene, 1 probe/300bp for 50kb flanking these genes, and 1 probe/500bp across recurrent microdeletions, as well as a backbone across the genome. The arrays were processed according to the manufacturer’s instructions and analysed with the Agilent Cytogenomics v.5.1.2.1 Software using the Default Analysis Method - CGH v.2. CNVs were filtered to exclude regions that 1) did not include a coding gene/exon (Ref. build Hg19); 2) were smaller than 10 kb; or 3) had a 50% overlap with CNVs detected in healthy published controls (excluding the recurrent CNV regions 15q11.2, 16p13.11, 16p11.2)(1).

*GNB1, MTOR, SLC2A1, KCNA2, CHRNB2, ASH1L, ATP1A2, CACNA1E, KCNH1, FBXO28, HNRNPU, LGI1, SLC25A22, KCNC1, SLC1A2, GRIN2B, SCN8A, FOXG1, DYNC1H1, UBE3A, GABRB3, CHRNA7, CHD2, NPRL3, TSC2, TBC1D24, RBFOX1, GRIN2A, STX1B, GNAO1, SLC13A5, PNPO, TCF4, HCN2, CACNA1A, SCN1B, GRIN2D, PNKP, MBD5, CACNB4, SCN3A, SCN2A, SCN1A, PLCB1, SNAP25, KCNB1, CHRNA4, KCNQ2, EEF1A2, DYRK1A, SIK1, DEPDC5, SLC6A1, NPRL2, FOXP1, TBL1XR1, GABRB1, HCN1, MEF2C, NR2F1, PURA, GABRB2, GABRA1, GABRG2, SYNGAP1, CHRNA2, KCNQ3, RORB, STXBP1, DNM1, TSC1, KCNT1, GRIN1, CLCN4, CDKL5, ARX, CASK, SYN1, SLC35A2, WDR45, IQSEC2, SMC1A, ARHGEF9, KIAA2022, BRWD3, PCDH19, DCX, MECP2, FLNA*

**Supplementary Note 4**: DEE starter panel design (32 genes)

Underlined: genes associated with neonatal/infantile onset epilepsy syndromes, with actionable implications

for treatment(2, 3).

*ALDH7A1, ARX, CACNA1A, CDKL5, CHD2, FOXG1, GABRA1, GABRG2, GRIN2A, GRIN2B, KCNA2, KCNB1, KCNQ2, KCNQ3, KCNT1, MECP2, PCDH19, PNPO, POLG, PRRT2, SCN1A, SCN1B, SCN2A, SCN8A, SCN9A, SLC25A22, SLC2A1, SLC6A1, STXBP1, SYNGAP1, TSC1, TSC2.*

***References***

1. MacDonald JR, Ziman R, Yuen RKC, Feuk L, Scherer SW. The Database of Genomic Variants: A curated collection of structural variation in the human genome [Internet]. *Nucleic Acids Res.* 2014;42(D1). doi:10.1093/nar/gkt958

2. Myers KA, Scheffer IE. Precision Medicine Approaches for Infantile-Onset Developmental and Epileptic Encephalopathies [Internet]. *https://doi.org/10.1146/annurev-pharmtox-052120-084449* 2022;62(1):641–662.

3. Bayat A, Bayat M, Rubboli G, Møller RS. Epilepsy Syndromes in the First Year of Life and Usefulness of Genetic Testing for Precision Therapy [Internet]2021;12(7):1051.
